# Retailers’ Perspectives on Stocking Healthy Food in Urban Poor Areas of Kuala Lumpur, Malaysia: A Qualitative Study

**DOI:** 10.64898/2026.09.24.26363967

**Authors:** Anis Munirah Mohd Sakri, Sameeha Mohd Jamil, Che Aniza Che Wel, Yong Kang Cheah, Huay Woon You, Adila Fahmida Saptari, Sirinya Phulkerd, Elaine Q Borazon, Bee Koon Poh, SEAOFE Study Group

## Abstract

High calorie intake of unhealthy foods are the main factor of overweight and obesity. In food environment, food retailers are the biggest player in shapping consumers’ purchase pattern. In this South East Asia Obesogenic Food Environment (SEAOFE) study, we aimed to to explore retailer perspectives on stocking decisions and barriers to stocking healthy food products in urban poor areas of Kuala Lumpur, Malaysia. The study was conducted using purposive sampling at three parliaments which have the highest urban poor population. 35 retailers who involved in procurement, retail operations, and product placement, with at least one year of experience, were selected as participants of this study. The interview were conducted face-to-face and been transcribed verbatim for data analysis. Nine key themes emerged regarding factors affecting stocking decisions: 1) consumer demand, 2) consumer satisfaction and feedback, 3) profitability, 4) forecast software, 5) fast-moving products, 6) product healthiness, 7) availability, 8) seasonality, and 9) e-commerce; while four key barriers in stocking healthy food products included 1) price, 2) shelf life, 3) consumer demand and preferences, and 4) availability of supply. This study showed that the urgency of targeted interventions to improve healthy food supply through collaboration from retailers, suppliers and policymakers. By addresing the issue will help to increase consumption of healthy foods and improve the health of the residents within the urban poor areas of Kuala Lumpur.

## Introduction

The current statistic showed that 1 in 2 adult is overweight and obese and it has become one of the main public health issues among Malaysian adults.(1) Similarly, abdominal obesity is also increasing to 54.5% in 2023(1). This serious issue is one of the major factors that contribute to non-communicable diseases (NCDs) including diabetes, hypertension, high cholesterol and obesity which are major risk factors for heart disease and stroke(1). The current data also showed that over half a million adults in Malaysia live with four NCDs because of the growing health and economic burden related to obesity and unhealthy dietary behaviours. This obesity issue is influenced by a few factors which are biological, behavioral, social and environmental factors (1). Among of all these factors, the food environment has become the main determinant of dietary behaviours and health outcomes. The food environment refers to the physical, economic, policy, and sociocultural surroundings that influence individuals’ food choices and nutritional status.(2) Food retail outlets, including supermarkets, grocery stores, convenience stores, and mini-markets, are important components of the community food environment because they determine the types of foods available and accessible to consumers.(3) Easy access to energy-dense, nutrient-poor, and ultra-processed foods may encourage unhealthy dietary behaviours and excessive calorie consumption, thereby contributing to the growing prevalence of overweight and obesity(4-5).

Previous studies have reported that the availability of unhealthy foods was greater than healthy foods in food retail, particularly in developed countries such as the United States, Australia and New Zealand (4-6). This environment is known as “food swamps” where unhealthy foods are more available compared to healthy foods. In one of the studies, the results indicated that accessing healthy food in retail outlets was quite challenging, especially in lowincome and non-urban areas where retailers had barriers to supply healthy foods, especially fresh fruits and vegetables due to logistic issues (7). This showed that people who live in the area are more exposed to unhealthy food environment. Hence, by increasing the availability of healthy foods within these urban poor areas represents a great strategy to promote healthier dietary intake and can also reduce the risk of obesity.

In conjuction with stocking healthy foods at retail stores, retailers are the main character in shaping healthy food environment because they are the ones who supply all the food products to retail stores. Through their stocking decisions, retailers influence which products should be available and accessible to consumers. Based on the previous study showed that retailers stocking decisions are influenced by many factors, including consumer demand, product selection, price and promotion(6). They always prioritize stocking products that are high in demand which also reflected consumer purchasing patterns on food availability and product placement within retail outlets (6). However, this study also reported that retailers in low socioeconomic areas often faced the challenge of stocking healthy foods due to limited purchasing power among consumers (6). The price of healthy products became the main issue for consumers. This challenge has discouraged retailers to supply healthy foods, despite the benefits to consumers.

While this factor has been extensively explored in high-income countries, such as United States, evidence from low- and middle-income countries, including Malaysia, remains limited. Most Malaysian studies examining food environments have primarily focused on consumer dietary behaviours, and not muchattention has been given to the perspectives of retailers who directly influence the local food supply. Furthermore, there is limited understanding of how retailers make their decisions to stock up products in urban poor areas, especially in the provision of healthier food options. This knowledge gap hinders the creation of effective interventions aimed at improving access to nutritious food, especially in urban poor areas. Therefore, this study aims to explore retailers’ perspective on stocking decisions and barriers to stocking healthy food products in urban poor areas of Kuala Lumpur, the capital city of Malaysia. Exploring how retailers make decisions about what they stock, when, and why, as well as knowing the extent to which they consider the healthiness of products they sell could potentially shed light in the current health issues and serve as a guideline for future policy formulation (5, 6).

## Methods

This study employed a qualitative design to explore the perspectives of food retailers regarding their stocking decisions and the barriers to providing healthy food products in urban poor areas of Kuala Lumpur. The study locations were determined based on three parliamentary areas with the highest percentage of urban poor population, namely Batu, Kepong and Bandar Tun Razak, based on data provided by Kuala Lumpur City Hall). Analytic Hierarchy Process (AHP) method was done to select the types of food retail outlets with experts from different fields, such as academia, Ministry of Health, food retailers, Malaysian Retailers Association (MRA), Federation of Malaysia Consumer Associations (FOMCA) and Kuala Lumpur City Hall. Each expert provided the assessment based on their expertise. They were asked to contribute in a short questionnaire which need themto prioritise the type of food retailers that were most relevant in urban poor communities. This method is commonly employed for decision making in food retail research when the situation was complex (7). Once the types of food retail outlets were selected, a geocoding method known as QGIS version 3.18 was employed to select the location and number of retailers within the chosen area (8). A distance of 1–4 km radius between retail outlets and residential areas was also considered when selecting the retail locations for this study (9).

A purposive sampling approach was used to recruit the participants, who were mainly retail personnel from modern and traditional retail formats located in the three selected areas. The inclusion criteria were retailers who played a major role in procurement, retail operations, and product placement, and who had at least one year of experience in their respective positions. For modern retailers, the main participants were store managers or operations managers, who were responsible for daily operations, human resources, marketing, promotion, pricing, and store display. Sales managers focusing on marketing, promotion, pricing, and store display strategies were also included in the study. For traditional retailers, store managers were recruited as study participants. In-depth interviews were conducted at the selected food retail locations from September 2022 to February 2023. This study is part of the South East Asian Obesogenic Food Environment (SEAOFE) project (10). Ethical approval was obtained from the university research ethics committee and permission to conduct the study was given by Kuala Lumpur City Council (DBKL). After selecting the food retailers, written consent forms were provided to the store managers of the chosen food retail outlets.

Interview questions were designed based on the discussion with local and international advisory groups who had expertise in food retail, food policy and research methods. The questions were employed to explore food retailers’ perspectives on stocking decisions and barriers to providing healthy food products. The interview guideline and questions were pretested prior to data collection to ensure the validity of the questions. Each interview lasted approximately 30-40 minutes per session. While the interviews were conducted face-to-face, anonymity of responses was assured.

Data saturation, indicating that no new themes emerged, was reached after interviewing the 35^th^ retailer. Each interview was audio-recorded and subsequently transcribed verbatim for data coding. The transcripts were thematically analyzed using Taguette software (11). The data was analyzed using the six steps of thematic analysis (12): (1) familiarization with the data; (2) generation of initial codes to describe the content; (3) searching for themes or patterns; (4) review of themes; (5) definition of themes; and (6) production of the report of the thematic analysis. Through this process, themes, linkages, and explanations were identified, and both deductive and inductive approaches were used in the analysis. First-order and second-order themes were developed to provide a better understanding of factors influencing food retailer decisions. The coding process was carried out by two coders, who reviewed and resolved inconsistencies in the codes. After completing the coding process, the codes were grouped into categories and themes.

Several approaches were employed to ensure the quality and trustworthiness of this study. Engagement with retailers were done to ensure that researchers established a strong rapport with participating retailers. The engagement was important to build trust and encourage the retailers to provide reliable data (13). Other than that, member checking was done to improve the accuracy of thematic analysis (14). A third individual, who was a postgraduate student, was involved in analysis process to verify interpretations and to improve the accuracy of findings.

A total of 35 retailers were recruited in this study, including 16 retailers from modern retail outlets (6 from hypermarkets, 6 from supermarkets, and 4 from convenience stores) and 19 from traditional stores (all of which were sundry shops).

## Results

Table 1 describes the summary of participants’ demographic profile and characteristics of the retail outlet. Majority of the retailers were male (74.3%). Most participants were store manager (88.6%). In terms of retail format, 45.7% represented modern retail outlets, while 54.3% were from traditional retail formats. The retail size varied with micro-sized being the largest group (54.3%) and small retailers were the smallest group (5.7%). Most of the ownership was local company, with 91.4% of business being locally owned.

**Table 1.** Characteristics of respondents (n = 35)

| Variable | Frequency | Percentage |
| --- | --- | --- |
| **Gender** |  |  |
| Male | 26 | 74.3 |
| Female | 9 | 25.7 |
| **Position in company** |  |  |
| Store Manager | 31 | 88.6 |
| Senior Division Manager & Operation Manager | 2 | 5.7 |
| Merchandiser & Buyer Manager | 2 | 5.7 |
| **Retail format** |  |  |
| Modern | 16 | 45.7 |
| Traditional | 19 | 54.3 |
| **Retail size** |  |  |
| Large | 8 | 22.9 |
| Medium | 6 | 17.1 |
| Small | 2 | 5.7 |
| Micro | 19 | 54.3 |
| **Ownership Type** |  |  |
| Local | 13 (Modern) | 37.1 |
|  | 19 (Traditional) | 54.3 |
| Foreign | 3 (Modern) | 8.6 |

### i) Stocking decisions for food and beverages in retail premises/stores

Figure 1 shows the 9 themes for under stocking decisions for food and beverages in retail premises, and Table 2 showed the each of the themes with quotes from participants.

**Table 2.** Themes for stocking decisions for food and beverages in retail premises/stores.

| Topic | Themes | Quotes |
| --- | --- | --- |
| Stocking decisions | Customer demand | *"We usually stock up based on customer request. We will try to fulfil their request. I will send the request to HQ to proceed with the order"* (Store manager, Supermarket 2).<br>*"I decided to restock the products based on customer's demand. I will sell what local customers use and want...."* (Store manager, Supermarket 6).<br>*"We stock up based on what customer demand here"* (Store manager, Traditional retailer 2). |
|  | Consumer satisfaction and feedback | *"We usually restock based on feedback and review from customers"* (Store manager, Traditional retailer 6).<br>*"We consider customer satisfaction before we order the stocks"* (Merchandiser & buyer manager, Hypermarket 2). |
|  | Profitability | *"We do consider about profit. If I take more stocks, I need to calculate the profit and need to utilize all the stocks"* (Store manager, Supermarket 6). |
|  | Forecast software | *"We use auto order for food products and daily use products. We forecast for extra order especially for fast moving items. We also have to forecast more especially during pay day"* (Store manager, Hypermarket 3). |
|  | Fast-moving products | *"We usually restock based on fast moving items"* (Store manager, Traditional retailer 8). |
|  | Healthiness of products | *"People consume honey and Chinese herbs more after COVID-19"* (Store manager, Supermarket 1).<br>*"People are more concerned about their health. For example, the demand for lemon and honey is more after the pandemic".* (Store manager, Traditional retailer 10). |
|  | Availability of products | *"We restock based on availability of products and we will also try to find the products until we get it"* (Store manager, Traditional retailer 8). |
|  | Seasonality | *"For festive season like Deepavali, we will standby laddu, sweet dessert. During CNY, we will prepare more food, and for Hari Raya we will prepare more ready-made ketupat"* (Store manager, Traditional retailer 9). |
|  | E-commerce | *"We restock based on comparison between last month's sales and current sales in both platforms, physical shop or online shop"* (Store manager, Supermarket 3). |

**Figure 1.**
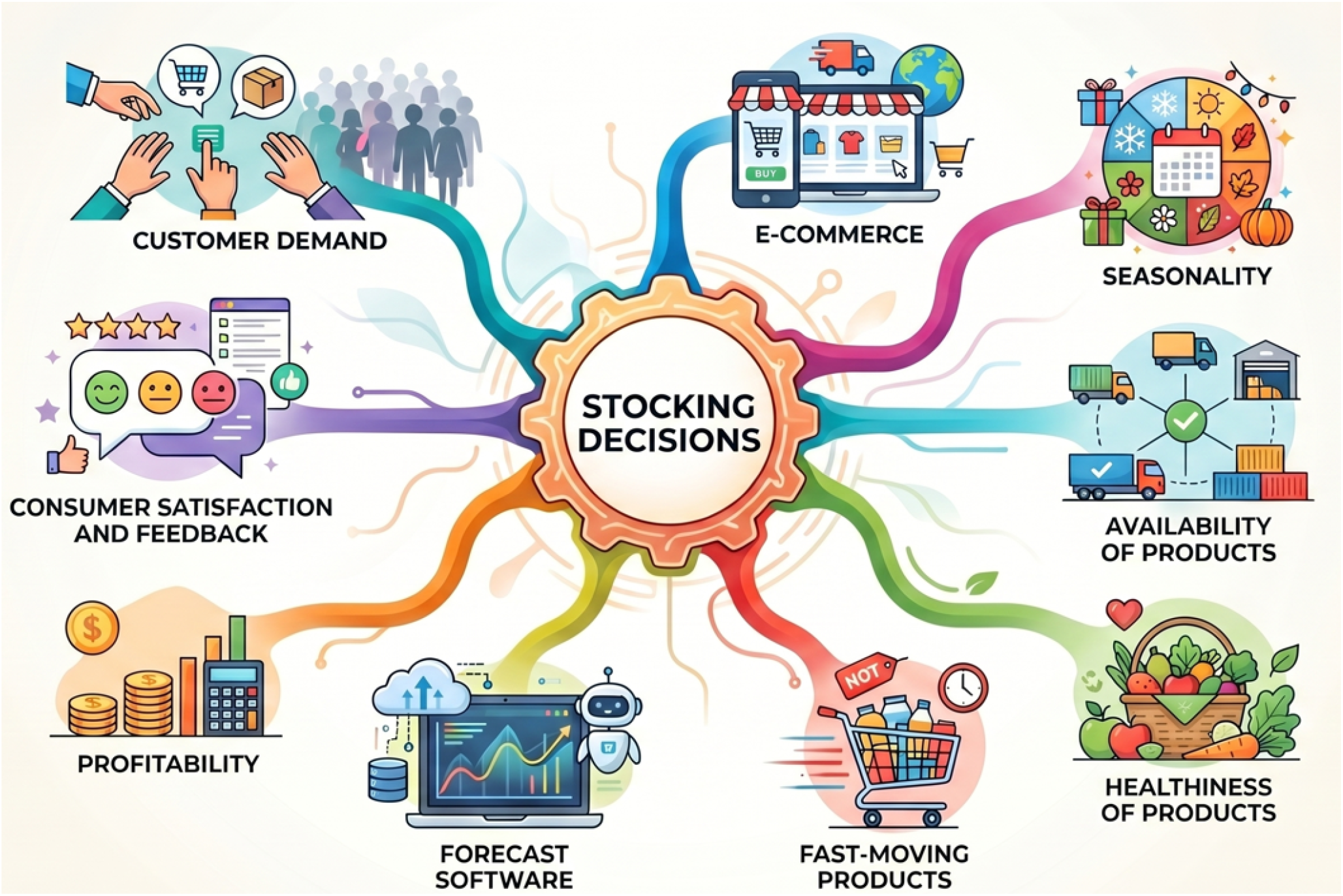
Themes for stocking decisions for food and beverages in retail premises/stores

### ii) Barriers to stocking healthy food and beverages in retail stores

Figure 2 shows the four key themes emerged on barriers to stocking healthy food and beverages in food retail premises and Table 3 showed the each of the themes with quotes from participants.

**Table 3.**
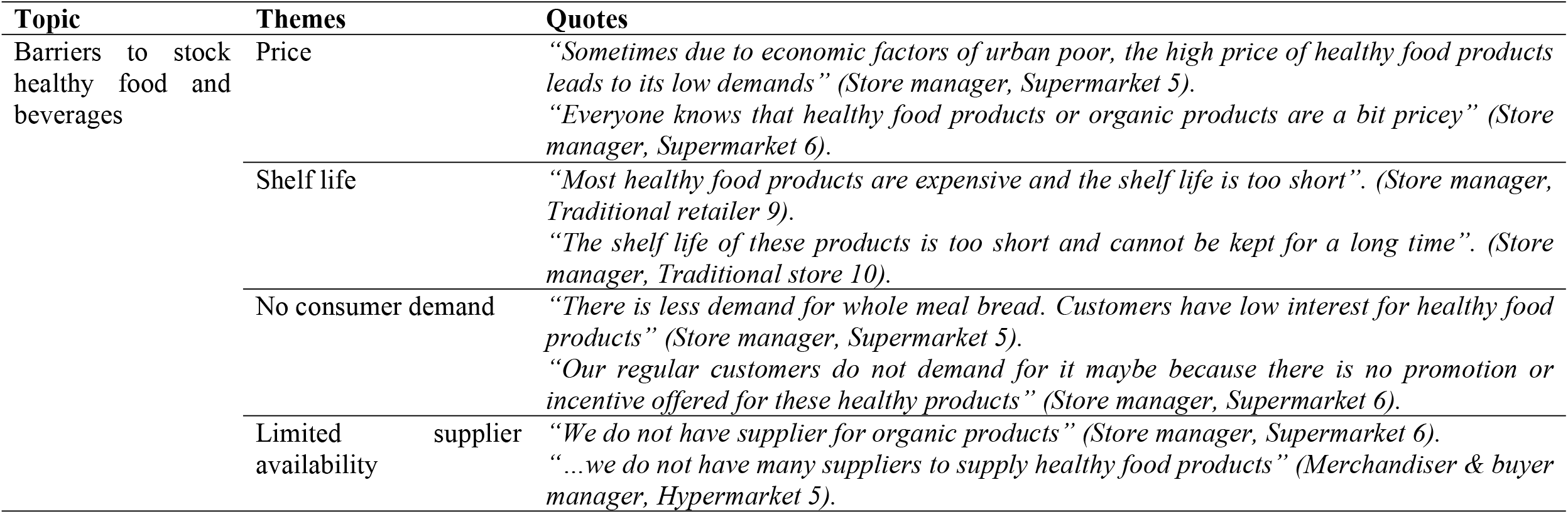
Themes for barriers to stock healthy food and beverages in retail premises/stores.

**Figure 2.**
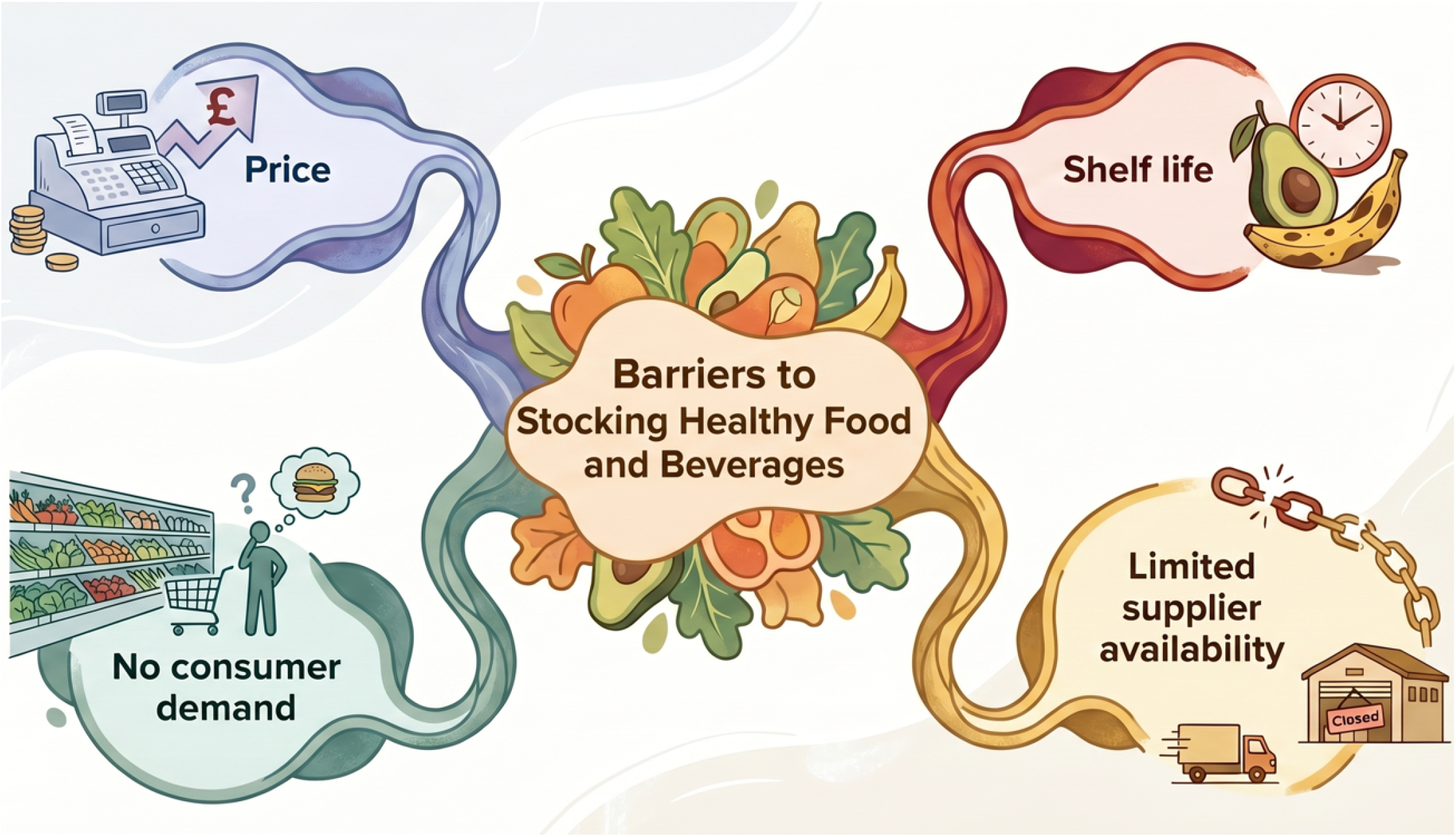
Themes for barriers to stock healthy food and beverages in retail premises/stores

## Discussion

This qualitative study explored retailers’ perspectives on food and beverage stocking decisions and examined the barriers to stocking healthier food products in urban poor communities in Kuala Lumpur, Malaysia. The findings suggest that retail stocking practices are shaped primarily by market and commercial considerations, with consumer demand and feedback playing a central role. Profitability, product movement, seasonality, availability, and perceived healthiness also influenced stocking decisions, while modern retailers additionally relied on forecasting systems to manage inventory. At the same time, retailers identified several interconnected barriers to stocking healthier products, particularly their relatively high prices, short shelf life, limited consumer demand, and inconsistent supply. Collectively, these findings illustrate that the availability of healthier foods in disadvantaged communities is not determined by consumer preferences alone, but emerges from the interaction between consumer demand, retailer economics, and supply-chain conditions. Consumer demand and feedback were consistently identified as important considerations when retailers decided which products to stock and replenish. Responsiveness to consumer preferences was considered essential for maintaining sales and sustaining business performance (15). This finding suggests that retailers operate within a demand-responsive environment in which product availability is closely linked to expected sales performance. Previous research involving 20 supermarket retailers in New York similarly found that consumer demand was an important determinant of product selection (6). Understanding and responding to consumer demand may therefore allow retailers to adjust their product assortment more effectively, while maintaining sales performance.

Profitability was another important consideration, particularly among modern retailers. Retailers described using forecasting software to estimate stock requirements based on sales trends, seasonal variations and consumer demand. Such systems enable retailers to predict future purchasing patterns and adjust inventory accordingly. Accurate forecasting can reduce both overstocking and stock shortages, while improving the likelihood that products are available when consumers seek them. Previous research similarly indicates that inventory forecasting can contribute to optimizing stock levels, improving consumer satisfaction and supporting business sustainability(15-17). Seasonal forecasting is particularly relevant for products with fluctuating demand, as inappropriate stock levels may increase either the risk of product shortages or financial losses associated with excess inventory (18).

These findings highlight an important distinction between the public health objective of increasing availability of healthy food products and the commercial realities of food retailing. From a public health perspective, increasing the number of healthy food products available in stores may appear straightforward. However, retailers must simultaneously consider product turnover, expected profitability, storage requirements, and the risk of unsold inventory. Consequently, interventions that seek to improve availability of healthy food products are more likely to be sustainable when retailers’ incentives and commercial constraints are incorporated into their design. Recent evidence on the long-term sustainability of healthy food retail interventions similarly emphasizes the importance of retailer-related and contextual determinants in maintaining changes over time.

Following the COVID-19 pandemic, retailers reported that consumers had become more health-conscious and increasingly interested in healthy food products associated with immune support and overall well-being. This finding is consistent with previous research documenting changes in food purchasing and consumption behaviors during and following the COVID-19 pandemic, including greater attention to health-related food choices (19). The pandemic may therefore have created an opportunity for retailers to respond to growing health consciousness by expanding the availability and visibility of healthier products.

Despite growing health consciousness among consumers, increased awareness does not necessarily translate into sustained changes in purchasing behavior. Retailers identified several persistent barriers to stocking healthy food products in urban poor communities. Product price was the most prominent concern. The findings indicate that healthy food products, particularly fresh fruits, vegetables, and organic products, were relatively expensive, potentially limiting their affordability among consumers in economically disadvantaged communities. From retailers’ perspective, limited consumer purchasing capacity increases the risk that healthy food products will remain unsold, resulting in financial losses and discouraging retailers from allocating shelf space to these products. This finding is consistent with previous qualitative research conducted among twenty retailers in the United States, where product price was identified as an important constraint on the stocking and sale of healthy food products (20).

The relationship between price, demand, and stocking practices may create a self-reinforcing cycle within disadvantaged food environments. When healthy food products are perceived as unaffordable, consumers may favor cheaper alternatives, which may often be energy-dense and less nutritious. Lower demand subsequently reduces retailers’ incentives to stock healthy food products, further limiting their availability within the local food environment (20). Thus, improving access to healthy foods in urban poor communities may require interventions that address both sides of the market rather than focusing exclusively on consumer education. Measures that reduce the relative price of healthy food products, increase their commercial attractiveness to retailers, or reduce the financial risks associated with stocking perishable healthy food products may be particularly relevant.

Product perishability represented another practical constraint. Retailers reported difficulties in managing products with relatively short shelf lives, particularly fresh fruits, vegetables, and other minimally processed or organic products. Compared with shelf-stable processed foods, these products may require more frequent replenishment and careful inventory management because deterioration can occur rapidly (21). The financial implications of product wastage may be particularly important for small retailers operating with limited resources. Previous research has similarly highlighted the importance of accurate demand forecasting to minimize spoilage of healthier perishable foods (22). In addition, appropriate storage facilities and efficient distribution systems are important for maintaining product quality and extending the period during which fresh products remain suitable for sale (23). Therefore, interventions aimed at improving the availability of healthier foods should consider not only product affordability but also the infrastructure and logistical requirements associated with their storage and distribution.

Limited consumer demand was also identified as a barrier to stocking healthy food products. Retailers reported that some consumers placed relatively little value on healthier food choices, particularly when these products were more expensive and received limited promotional support. This finding is consistent with previous evidence indicating that lower-priced, less healthy food products often attract greater consumer demand than healthier alternatives, particularly in lower-income settings (24). The absence of effective promotion may further reduce consumers’ exposure to healthier options and their willingness to purchase them.

The findings also suggest that local food-related initiatives may influence consumer demand for healthy food products and purchasing patterns within some urban poor communities. For example, programs such as urban farming and MyGrocer in selective urban poor flats (PPR) communities under initiatives implemented by the city council (DBKL) may provide residents with alternative sources of vegetables and other food products. These initiatives are intended to strengthen food security and promote community self-sufficiency, potentially reducing reliance on conventional retail outlets for selected healthy food products . Nevertheless, this interpretation remains speculative based on the present data and should be examined in future research. Understanding how community-based food initiatives interact with formal and informal retail environments would provide valuable insight into the broader food environment of urban poor communities.

In addition to demand-related constraints, retailers described difficulties in obtaining a reliable supply of healthier products. Limited supplier availability may restrict retailers’ ability to introduce or maintain healthier products, even when there is an interest in doing so. An inconsistent supply can also make it difficult for retailers to ensure continuous product availability and may increase the perceived commercial risk of stocking these items. Previous research has emphasized the importance of strengthening the supply of healthier products within retail environments, as improved availability can provide consumers with greater opportunities to purchase healthier foods and potentially stimulate demand (24). This highlights the need to consider the food supply chain as part of efforts to improve the healthfulness of retail food environments. Collaboration among retailers, wholesalers, distributors, and policymakers may be necessary to develop more reliable supply channels for healthier products in underserved communities.

Taken together, the findings indicate that the availability of healthier foods in urban poor retail environments is shaped by a combination of consumer, commercial, and supply-side factors. Retailers operate within a market in which consumer demand and profitability are closely connected to stocking decisions. Consequently, simply encouraging retailers to stock healthier products may have limited impact if the underlying economic and logistical constraints remain unresolved. A more comprehensive approach should therefore seek to increase consumer demand while simultaneously reducing the perceived financial and operational risks associated with stocking healthier products.

Several limitations should be considered when interpreting the findings. First, the study was conducted exclusively in urban poor communities in Kuala Lumpur, and only three of the 11 parliamentary constituencies were included. Therefore, the findings may not represent all areas in Malaysia, particularly those with different socioeconomic, geographical, or retail characteristics. Second, some retailers provided relatively brief responses during the interviews, which limited the depth of information that could be obtained for certain topics. Differences in the extent of participants’ engagement may also have influenced the richness of the qualitative data.

Nevertheless, this study has several strengths. The inclusion of both modern and traditional food retailers provided perspectives from different types of retail settings and allowed a broader understanding of stocking practices within urban poor communities. To the best of our knowledge, this study is among the few studies in Malaysia to examine retailers’ perspectives across both modern and traditional food retail sectors in relation to the availability of healthy food products. Previous studies in Malaysia predominantly examined food purchasing from the consumer perspective, leaving relatively limited evidence concerning the role of retailers in shaping the local food environment. By examining retailers’ stocking decisions alongside the barriers they encounter, this study provides insights into an important but comparatively understudied component of the food environment.

The findings have potential implications for policy and practice. Understanding the commercial realities faced by retailers may assist policymakers in developing interventions that are feasible within existing retail settings. Rather than placing responsibility solely on retailers or consumers, strategies should address affordability, supply reliability, product perishability, and consumer demand concurrently. Such an approach may strengthen the potential for healthier products to become more consistently available within disadvantaged communities.

## Conclusion

This study demonstrates that food stocking decisions among retailers in urban poor communities are strongly influenced by consumer demand and satisfaction, with profitability, product movement, perceived healthiness, product availability, and seasonality also contributing to these decisions. Modern retailers further relied on forecasting systems to anticipate demand and manage inventory. These findings highlight that food retail decisions are embedded within broader economic and supply-chain considerations rather than being determined by nutritional considerations alone.

The availability of healthier foods was constrained by several interconnected barriers, particularly relatively high prices, limited consumer demand, short shelf life, and difficulties in securing a consistent supply. These barriers may reinforce one another, creating a retail environment in which inexpensive, shelf-stable, and fast-moving products are commercially more attractive than healthier alternatives. Addressing these challenges therefore requires coordinated action across both demand and supply sides of the food system.

Strategies to improve the affordability and attractiveness of healthy food products should be complemented by measures that reduce retailers’ operational and financial risks, including improved supply networks, inventory management, storage facilities, and targeted retailer incentives. Collaboration among policymakers, retailers, suppliers, food manufacturers, and communities will be important for developing interventions that are both commercially feasible and aligned with public health objectives. Efforts to improve product availability, pricing, promotion, and accessibility may ultimately contribute to a more supportive food environment and provide urban poor communities with greater opportunities to make healthier food choices.

By incorporating retailers’ perspectives, this study adds to the limited evidence on the supply-side determinants of healthy food availability in Malaysian urban poor communities. These findings may inform future interventions and policies aimed at creating healthier and more equitable retail food environments in Malaysia, thereby supporting broader efforts to promote health and well-being in line with Sustainable Development Goal 3.

## Data Availability

-

## Acknowledgement

We are grateful to all retailers and researchers who contributed to this study.

## References

1. Health IfP. Non-Communicable Disease and Healthcare Demand National Health & Morbidity Survey 2023 In: (NIH) NIoH, editor. Malaysia: Institute for Public Health; 2024.

2. Turner C, Aggarwal A, Walls H, Herforth A, Drewnowski A, Coates J, et al. Concepts and critical perspectives for food environment research: A global framework with implications for action in low- and middle-income countries. Global Food Security. 2018;18:93–101.

3. Downs S, Glass S, Linn KK, Fanzo J. The interface between consumers and their food environment in Myanmar: an exploratory mixed-methods study. Public Health Nutrition. 2018;22:1075–88.

4. Schultz S, Cameron AJ, Grigsby-Duffy L, Robinson E, Marshall J, Orellana L, et al. Availability and placement of healthy and discretionary food in Australian supermarkets by chain and level of socio-economic disadvantage. Public Health Nutr. 2021;24(2):203–14.

5. Gittelsohn J, Kasprzak CM, Hill AB, Sundermeir SM, Laska MN, Dombrowski RD, et al. Increasing Healthy Food Access for Low-Income Communities: Protocol of the Healthy Community Stores Case Study Project. Int J Environ Res Public Health. 2022;19(2).

6. Martinez O, Rodriguez N, Mercurio A, Bragg M, Elbel B. Supermarket retailers’ perspectives on healthy food retail strategies: in-depth interviews. BMC Public Health. 2018;18(1):1019.

7. Lipovetsky S. Understanding the Analytic Hierarchy Process. Technometrics. 2021;63(2):278–9.

8. Team QD. QGIS 2020 [Available from: https://qgis.org/.

9. Mahendra A, Polsky JY, Robitaille É, Lefebvre M, McBrien T, Minaker LM. Status report - Geographic retail food environment measures for use in public health. Health promotion and chronic disease prevention in Canada : research, policy and practice. 2017;37 10:357–62.

10. Phulkerd S, Rachmi CN, Sameeha MJ, Borazon EQ, Thow AM, Trevena H, et al. Identifying Opportunities for Strategic Policy Design to Address the Double Burden of Malnutrition through Healthier Retail Food: Protocol for South East Asia Obesogenic Food Environment (SEAOFE) Study. Int J Environ Res Public Health. 2022;19(1).

11. Rampin RRaV. Taguette: open-source qualitative data analysis. Journal of Open Source Software. 2021; 6(68)(3522).

12. Clarke V, Braun V. Thematic analysis. The Journal of Positive Psychology. 2016;12:1–2.

13. Hadi MA, José Closs S. Ensuring rigour and trustworthiness of qualitative research in clinical pharmacy. International Journal of Clinical Pharmacy. 2016;38(3):641–6.

14. Chen H, Ellett JK, Phillips R, Feng Y. Small-scale produce growers’ barriers and motivators to value-added business: Food safety and beyond. Food Control. 2021;130:108192.

15. Caspi CE, Pelletier JE, Harnack L, Erickson DJ, Laska MN. Differences in healthy food supply and stocking practices between small grocery stores, gas-marts, pharmacies and dollar stores. Public Health Nutr. 2016;19(3):540–7.

16. Gravlee CC, Boston PQ, Mitchell MM, Schultz AF, Betterley C. Food store owners’ and managers’ perspectives on the food environment: an exploratory mixed-methods study. BMC Public Health. 2014;14:1031.

17. Tadayonrad Y, Ndiaye AB. A new key performance indicator model for demand forecasting in inventory management considering supply chain reliability and seasonality. Supply Chain Analytics. 2023;3:100026.

18. Ye L, Xie N, Boylan JE, Shang Z. Forecasting seasonal demand for retail: A Fourier time-varying grey model. International Journal of Forecasting. 2024.

19. Li Z, Zhao A, Li J, Ke Y, Huo S, Ma Y. Food and Nutrition Related Concerns Post Lockdown during COVID-19 Pandemic and Their Association with Dietary Behaviors. Foods. 2021;10(11).

20. Odoms-Young A, Brown AGM, Agurs-Collins T, Glanz K. Food Insecurity, Neighborhood Food Environment, and Health Disparities: State of the Science, Research Gaps and Opportunities. The American Journal of Clinical Nutrition. 2024;119(3):850–61.

21. Bojei J. Essential Quality Attributes in Fresh Produce Purchase by Malaysian Consumers. Journal of Agribusiness Marketing. 2010;3:1–19.

22. Wang L, Teplitski M. Microbiological food safety considerations in shelf-life extension of fresh fruits and vegetables. Current Opinion in Biotechnology. 2023;80:102895.

23. Abbas H, Zhao L, Gong X, Faiz N. The perishable products case to achieve sustainable food quality and safety goals implementing on-field sustainable supply chain model. Socio-Economic Planning Sciences. 2023;87:101562.

24. Haboush-Deloye AL, Knight MA, Bungum N, Spendlove S. Healthy Foods in Convenience Stores: Benefits, Barriers, and Best Practices. Health Promotion Practice. 2023;24(1_suppl):108S–11S.

